# Cost-benefit of Influenza Vaccinations in Frontline Workers: A Dynamic Modelling Study

**DOI:** 10.1101/2024.08.27.24312639

**Authors:** Peter Mortensen, William Gilks, Selina Kim, Richard Bennett, Matthew Linley

**Affiliations:** Airfinity Ltd, London, United Kingdom

## Abstract

Influenza significantly impacts public health, particularly among the elderly and those with underlying health conditions, but it also imposes substantial economic and operational burdens on the working-age population. This study introduces a novel machine learning-based Susceptible-Infected-Recovered (SIR) model solved as an agent-based model (ABM), designed to dynamically simulate influenza spread and assess the cost-benefit of vaccination programs specifically for frontline workers. Unlike traditional models, our approach accounts for the diverse contact rates and risk profiles across different job types, offering a more granular and accurate prediction of influenza’s impact on workforce productivity. We utilised historical influenza data from the CDC and WHO/FluMart to model the effects of varying vaccination coverage levels on infections, sick days, and associated costs within a typical workplace. The results demonstrate that higher vaccination coverage significantly reduces both the total number of infections and the peak sickness levels, leading to substantial cost savings. Additionally, higher vaccination coverage was associated with a significantly lower peak in sickness, mitigating periods of high absenteeism and operational disruptions. The model highlights the economic advantages of vaccination programs, particularly for sectors with higher salaries and absenteeism rates. It also underscores the importance of targeting frontline workers, who have higher contact rates and contribute more significantly to influenza transmission. This model’s ability to capture the dynamic nature of influenza transmission and its differential effects on various work types represents a significant advancement over previous static models. It provides a robust tool for organisations to optimise vaccination strategies, ensuring business continuity and enhancing productivity during influenza seasons.

## Introduction

Influenza is a significant public health concern, often highlighted for its severe impact on the elderly and those with underlying health conditions. The substantial morbidity and mortality associated with influenza in these vulnerable populations have led to a robust focus on vaccination strategies and healthcare resources aimed at mitigating these severe outcomes [1] [2]. However, there is a growing recognition of the substantial economic and operational impact that influenza can impose on the working-age population, who, despite generally experiencing less severe disease outcomes, still contribute to the overall burden of the illness.

While younger, healthier individuals are less likely to be hospitalised due to influenza, they are still susceptible to contracting the virus. Influenza can lead to significant absenteeism from work, with employees potentially missing several days due to illness [3] [4] [5]. This absenteeism not only affects individual productivity but can also impose considerable costs on organisations and the broader economy. The financial implications extend beyond direct medical costs to include lost productivity, operational disruptions, and increased workloads for remaining employees [5].

The economic burden of influenza in the working-age population has been increasingly recognised as a critical factor in the cost-benefit analysis of vaccination programs. Research has shown that influenza-related absenteeism can lead to substantial economic losses for both private and public sector organisations [6] [7]. These costs underscore the potential economic advantages of investing in vaccination programs targeted at this demographic, which may reduce absenteeism and associated costs, thereby benefiting organisations and the economy at large.

Previous studies have demonstrated the value of influenza vaccination from various perspectives, including direct medical cost savings and indirect benefits such as reduced absenteeism and enhanced productivity. For instance, a review by Nichol [8] showed that vaccination programs could result in significant economic savings by reducing both healthcare expenditures and productivity losses. Other studies, such as that by Campbell et al. [9], have also highlighted the cost-effectiveness of workplace vaccination programs in preventing influenza and reducing associated economic burdens. However, many of these previously reported studies have used simplistic techniques for estimating the benefits of vaccination, and do not accurately capture the dynamic nature of the infectious disease spread in different working groups and across age demographics. Specifically, the additional benefits that could be seen in frontline workers, which have higher contact rates and are often critical workers, are not accurately accounted for in current methodologies.

In this study, we aim to address this gap by developing a novel method to assess the cost-benefit of influenza vaccination specifically for the frontline working population. Here we develop a dynamic machine-learning compartmental model coupled with an agent-based model (ABM) to reflect the dynamic nature of influenza and how it affects outcomes in different working populations. We consider differences in working types between frontline workers and office staff, where these differences have been shown to significantly impact contact rates [10]. Our approach evaluates the economic advantages of vaccination from the perspective of organisations, considering both direct and indirect costs. By providing a detailed analysis of the potential financial benefits associated with workplace vaccination programs, we hope to contribute valuable insights that can inform policy decisions and organisational strategies regarding influenza vaccination.

## Methodology

### Data Collection and Processing

To evaluate the cost-benefit of influenza vaccination in the working population, we collected historical age-stratified influenza data from various sources. Weekly new cases data, season totals for hospitalisations and deaths were obtained from the Center of Disease Control and Prevention [11]. Specifically, estimates of the true number of infections, hospitalisations, and deaths by age group and influenza season were accessed from the CDC’s ‘Burden of Flu’ website [12]. These data were utilised to calculate infection-to-hospitalisation and infection-to-death rates for each season and age group.

To obtain values for the true number of new infections per week, the reported infections per week were expressed as a proportion of the total reported infections for the season, and then divided by the CDC’s estimated true infections for that season [12]. We separated the time series data into individual seasons from 2010 to 2020, excluding seasons after the arrival of COVID-19 lockdown measures in 2021 and 2022 as they were not considered standard. The age groups considered in this analysis were 18-49, 50-64, 65+, and an ‘All Ages’ group. The data were cleaned and processed to ensure accuracy before being utilised in the model.

Weekly reported infections data were obtained from the WHO [13], using ‘non-Sentinel’ and ‘Not Defined’ sources, which were preferred for the United States as they provide data over longer periods and at higher volumes compared to Sentinel sources. The typical low point of the influenza season was assumed to be calendar week 31, which was used to set the start and end dates for each season.

### Machine Learning SIR (Susceptible, Infected, Recovered) Model

To model the impact of workplace vaccinations, we employed a machine learning-based Susceptible-Infected-Recovered (SIR) model, incorporating additional compartments to better capture the dynamics of influenza spread. The model used here was adapted from our previous work for forecasting COVID-19 where it is described in more detail [14]. This model includes six primary compartments: susceptible (*S*), exposed (*E*), infectious (*I*), hospitalised (*H*), recovered (*R*), and deceased (*D*). Additionally, three non-conventional compartments—cumulative infections (*I*_*2*_), cumulative hospitalisations (*H*_*2*_), and hospital discharges (*L*)—were included to provide detailed estimates of daily new cases, hospitalisations, and discharges.

The ordinary differential equations (ODEs) for this model are defined as:

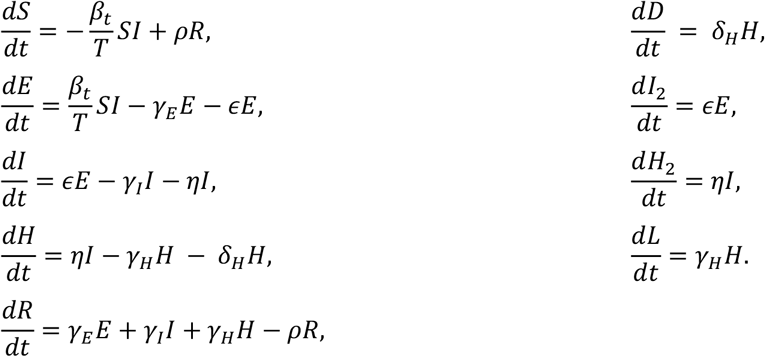

Here, *β*_*t*_, represents the time-varying infection rate, *ϵ*, is the rate of becoming infectious, *γ*_*E*_, *γ*_*I*_, and *γ*_*H*_, are the rates of recovering of the exposed group, the infectious group, and the hospitalised group, respectively, *η*, is the rate of, *ρ*, hospitalisation the rate of becoming susceptible again and, *d*, is the death rate.

This SIR model can be solved either as an equation-based model (EBM) or as an agent-based model (ABM). Solving the model as an EBM is fast and very efficient, but it assumes the population is entirely homogenous and well-mixed, and thus it does not allow for individuals to have their demographics impact their disease progression. However, an agent-based model will allow the population to have age-dependent parameters and, in the case of this work, job-dependent parameters. To establish these parameters, the previously established parameter fitting process was applied.

### Parameter Fitting

To determine the model parameters, we used the least-squares fitting algorithm provided by scipy.optimize.least_squares (SciPy). This algorithm finds a local minimum of a cost function by minimising the residual between model outputs and observed data. The residuals were computed using data on new cases, new hospitalisations, new deaths, and patient counts and the equivalent results of the SIR model, solved as an EBM. Time-varying parameters were employed to account for changes in disease dynamics over time, particularly the infection rate which decreases as the epidemic progresses and more individuals recover.

For each age group and season, parameters were initially estimated with the infection rate varying over time. Subsequently, we fixed the progression rates (*ϵ* and η) and used the previously estimated infection rates to fit seasonal values for recovery rates, hospitalisation rates, and death rates. These parameters were then compared with the vaccination coverage data to establish relationships between vaccination rates and disease outcomes.

This process produced three parameter sets for the three age-groups of interest and established the average change in the infection rate over the course of a year, which is identical for each age group. It should be noted that to perform this parameter fitting the SIR model was being solved as an EBM, thus assumed homogenous populations that are entirely in the given age-group. To apply the model to an ABM it assumed the difference between the age-specific parameters in a homogenous population and that of a population with a range of age groups, is negligible.

### Vaccination-dependent Parameters

Vaccine coverage has a significant impact on overall cases of influenza and the severity of outcomes [15]. In this work we were particularly interested in how the costs associated with employees taking time off due to influenza sickness can be mitigated by vaccinating the workforce. The parameter fitting process can find relationships between the estimated parameters from a given year and the associated vaccine coverage. Here it was assumed there is a linear relationship between the parameters and the vaccine coverage, which was then extrapolated out to find the associated parameter for a given vaccine level. However, these companies do not exist in a vacuum and have frequent interaction with the general public, which will not be impacted by a company’s particular vaccination campaign. In this study we assumed that the public remains constant in each scenario and the companies will interact with a number of customers relative to the company’s size. We assume that the vaccination coverage of the customers, *Vac*_*cust*_, is approximately 47% [16] (this is the average for the United States which will be the focus of the case study presented in this paper). In the following equation the vaccine coverage in the customer population is referred to as *Pop*_*cust*_, with vaccination rate in the company population shown as *Pop*_*comp*_. Thus, for a given company vaccination coverage the overall population vaccination coverage is:

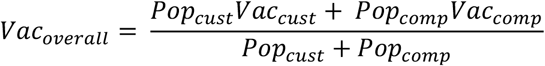

Using this scaling, when the company has a vaccine coverage of 0%, the overall vaccine coverage is approximately 31% and respectively, at the extreme end, when the company has a vaccine coverage of 80%, the overall vaccine coverage is approximately 58%.

### Agent-Based Model Implementation for Forecasting Scenarios

An agent-based model (ABM) approach was then used to solve the SIR model with a population with age-specific differences seen in influenza disease impact as well as changes in the contact rate for different job types. The SIR model parameters were converted to probabilities for use in an agent-based model. The conversion formula used was:

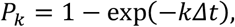

Where *P*_*k*_ is the probability of an agent making a transition for channel with the respective parameter *k* from the SIR model and Δ*t* is the time step. This approximation holds for suitably small enough time steps. For this model it was found that Δ*t* = 0.1 was sufficient. Below are comparisons between the SIR model being solved with as an ABM and an equation-based model (EBM). In this example the scenario has a homogenous population of 50,000 people, all 18–49-year-old front-line workers. The ABM was run 10 times with Δ*t* = 0.1, from which the mean of the outputs has been taken.

### Scenario Modelling

Using this established agent-based model a range of scenarios were generated based on known demographics of companies. Where each company can have varying number of employees, country locations, proportions of one three jobs and different age-stratification. Each agent in the population is within one of three age bands (18-49 years, 50-64 years and 65+ years) and one of three jobs (front line, back office and management). The front-line workers will have regular interactions with the general public through sales and services, whereas back office and management will tend to work in offices and have reduced interaction with the general public on a day to day basis, thus are assumed to have a 20% reduction in the fitted contact rate range (estimated from [10]). Although the contact rate is not an explicit parameter the infection rate, *β*_*t*_, can be assumed to be the equivalent to the product of the reproduction number and the contact rate. Thus, the employees who work in offices can have an adjusted contact rate, and thus an adjusted infection rate, to account for the increased sheltering received from not having as much direct contact with the general population.

The agent-based model was run for various vaccination scenarios: 0%, 20%, 40%, 60%, and 80% vaccine coverage to forecast scenarios for a given workplace. This simulation assessed the impact of different vaccination rates on influenza dynamics within the workforce.

The initial timings of the forecasted scenarios were fixed based on predicted wave peak timings. To predict wave peaks for each season, we used the most recent ten years of weekly data, excluding data from the 2021 and 2022 seasons due to the impact of COVID-19. The week-of-the-season was recalculated within each season so that the peak of infections was centred over week 26. This data was used as input for a linear model, with the season week as a categorical variable, to predict the wave shape relative to the season total. Smoothing was applied using a rolling mean with a window of three weeks. The predicted peak was identified as the equivalent calendar week with the maximum expected infections for that season. After identifying the expected peak week, the wave start date was determined from the curve and used as the initial timepoint for the SIR-ABM forecast scenarios.

### Cost-Benefit Analysis

For each simulation scenario, we calculated the expected number of sick days using the average sick days reported in the literature (three days for the US) [17]. We then estimated the cost of sick days by assuming that each sick employee is replaced by a temporary hire at an equivalent daily rate. The savings from vaccination were computed by comparing the costs associated with different vaccination rates against the baseline (0% vaccine coverage), including the costs for vaccinating the population and the savings achieved from fewer sick days taken.

### Assumptions and limitations

In our analysis, we made several key assumptions to ensure the feasibility and relevance of the model. We assumed that office workers, comprising those in back-office and management roles, experience a 20% reduction in the infection rate due to the nature of their jobs. This adjustment reflects the lower exposure risk associated with these positions. Vaccination is assumed to be administered before the onset of the influenza season, ensuring maximum effectiveness and that each vaccine costs $13.10. Here it was also assumed that the company covers all employee vaccinations, and no employee gets a vaccination of their own volition. However, as discussed previously, it was assumed that the overall vaccine coverage of the system is impacted by the general public and it was assumed that the company employees interact with twice as many customers as there are employees at the company. We also assumed a linear relationship between the percentage of vaccines administered and changes in model parameters, valid within the overall vaccine coverage. The model operated under the premise that the population is well-mixed. Consequently, the parameters derived from age-specific data were applied uniformly to the mixed population within the agent-based model. Employees were considered to work 8 hours a day, 5 days a week, and 48 weeks a year. If an employee falls ill, their absence was fully covered by a temporary hire who is paid an equivalent daily rate, but this temporary hire does not participate in the infection dynamics of the model.

### Case Study

We developed a case study of a hypothetical US-based company with 30,000 employees, with the workforce distributed to be: 70% front-line workers, 25% back-office workers, and 5% management. These job types each have distinct age-stratifications (as seen in the table below) and overall, the workforce is approximately 70% aged 18-49, 25% aged 50-64, and 5% aged 65+. Average annual salaries are $45,000 for general employees and $75,000 for management. Sick leave data indicates an average of three days off due to influenza [17]. This case study was used to apply the model findings and conduct the cost-benefit analysis of different vaccination coverage scenarios.

### Sensitivity analysis

Sensitivity analysis was performed to understand key parameters in the model. The first three variables tested were the company salary, the cost of the vaccines and the average number of days off. Here, a baseline case was run for a hypothetical company, with a size of 30,000 employees, with the job proportions being 70%, 25%, and 5% for the front-line worker, back-office workers and the management, respectively, with the three age stratifications being 70%, 25%, and 5% for the 18-49 year olds, 50-64 year olds and the those who are 65+, respectively. Each variable was varied one at a time, with their baseline values being an average company salary of $45,000, a single vaccination costing $13.08 and the average days off being 3 days, at a 47% vaccination coverage to reflect the average flu vaccination level in the US.

The second piece of sensitivity analysis fixed the company salaries, vaccine costs and the average days off, as per the baseline, and varied the proportion of the company that were frontline workers. This was completed for three vaccination coverage levels (0%, 40% and 80%), each with 100 simulations. The frontline worker proportion was randomly varied with a normal distribution, varying between 40% and 100%, with a mean of 70%, the remaining proportion of the 30,000 workers was divided with a constant 5:1 ratio between the back-office workers and managers, respectively. A histogram of the respective job type distributions is shown in Figure 3.

**Figure 1.**
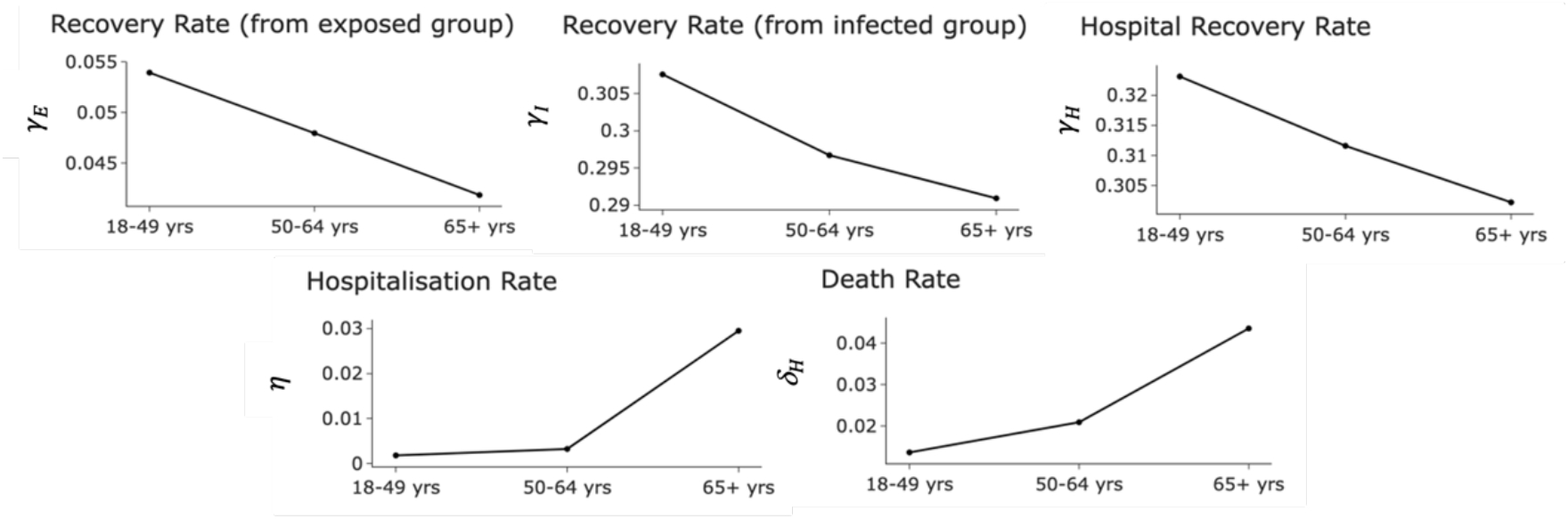
Plots of the variation in the machine learning-solved SIR model parameters with respect to the age group.

**Figure 2.**
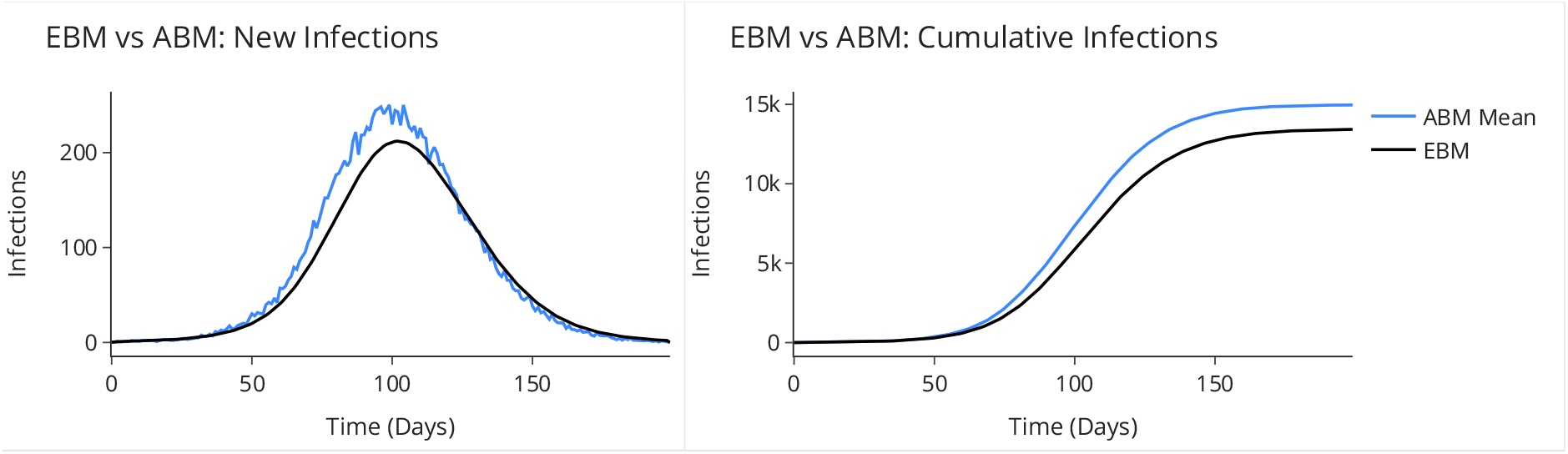
Plots comparing the results of the equation-based model and the agent-based model for new infections (left) and cumulative infections (right).

**Figure 3.**
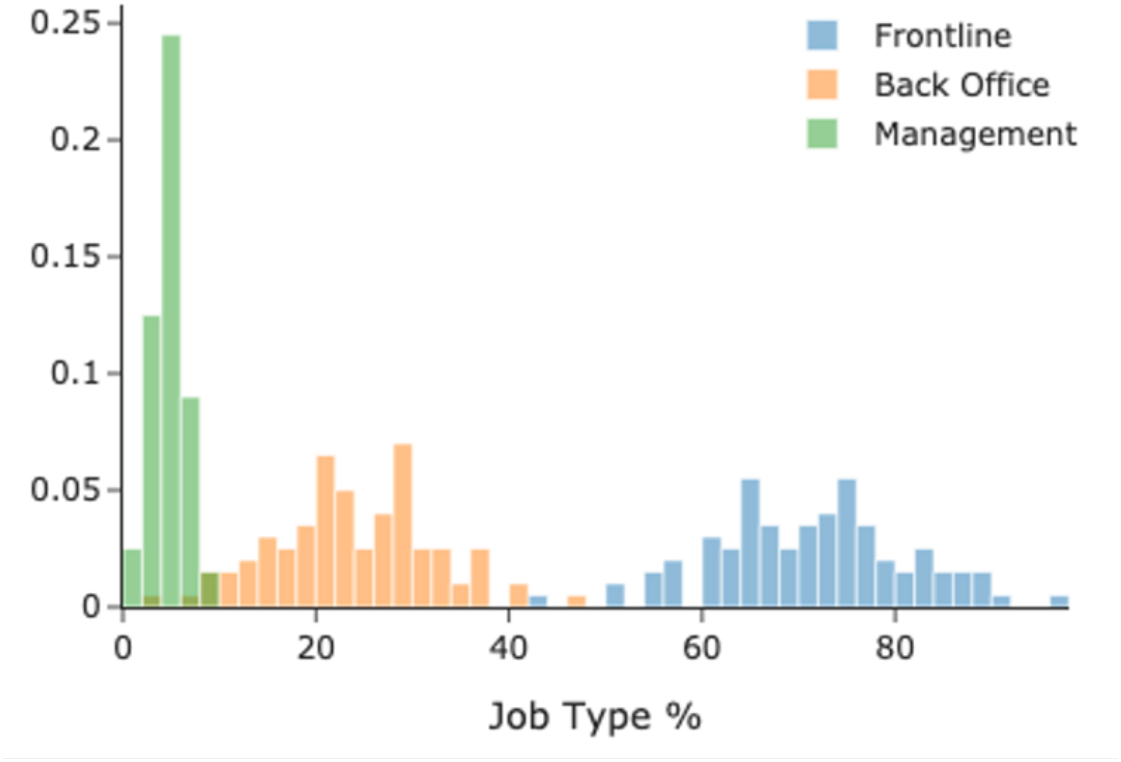
A histogram of the proportions of the three job groups in the 100 hypothetical companies of the sensitivity analysis with a vaccination coverage of 40%.

**Figure 4.**
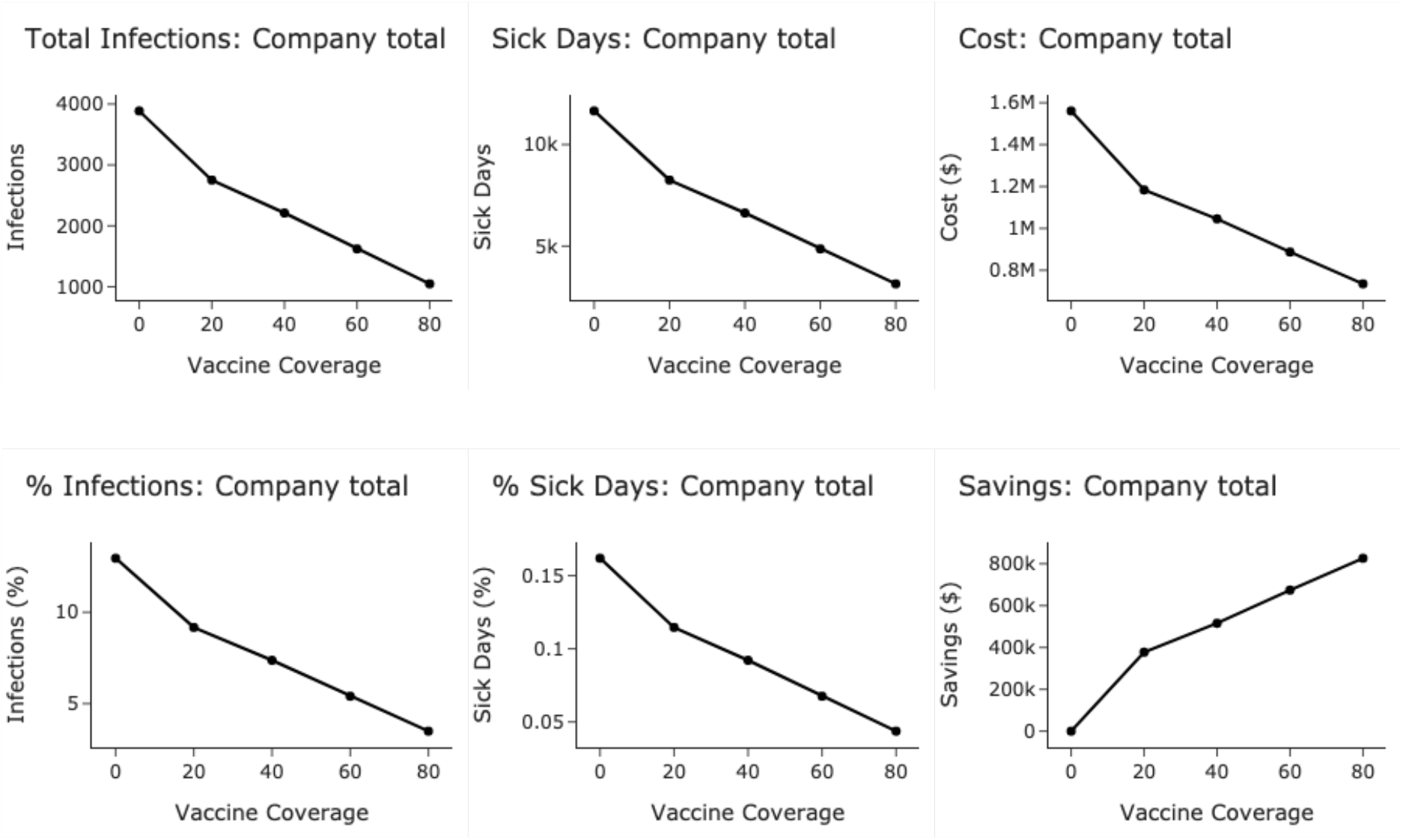
Plots showing outcomes for different metrics across different vaccine coverage scenarios from 0% to 80% vaccine coverage.

**Figure 5.**
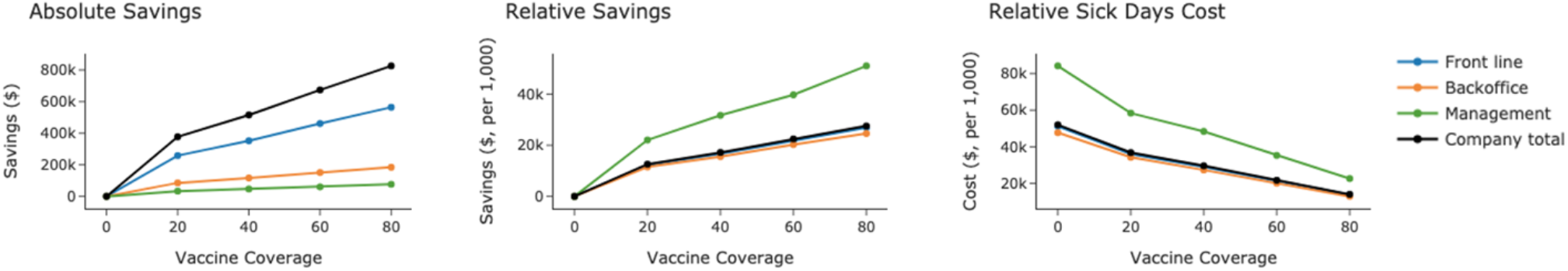
Plots showing outcomes for different metrics for each work banding, as well as company totals across different vaccine coverage scenarios from 0% to 80% vaccine coverage. Middle and right-hand plots show values per 1,000 people within each work banding.

## Results

The financial impact of influenza vaccinations on the hypothetical company was assessed across different work types and vaccination coverage scenarios. The modelled output of total infections and number of sick days are visualised as absolute numbers and as a percentage of the total for different vaccine coverages. When looking at the percentage, the total infections across the influenza season in the company are estimated to be 10% of the company with 0% vaccinations, which drops to 6% of the company getting infected at 80% vaccine coverage. The percentage of sick days is considerably lower, starting at 0.12% of all working days at 0% vaccine coverage, diminishing to less than 0.08% sick days at 80% vaccine coverage. Sick days are significantly lower than total infections due to sickness lasting for only a few days compared to the total number of days throughout the influenza season. Total costs due to sickness and vaccinations dropped with increasing vaccine coverage, with total costs at 80% vaccine coverage estimated to be approximately $735K compared with the $1.56M costs at 0% vaccine coverage. As a result, the cost savings were calculated to be just over $825K with the greatest vaccine coverage. The trend shows that there are increased savings with each additional vaccine administered.

The total savings achieved under varying vaccination rates are summarised in Table 2.

**Table 1:** Proportion of each age group in each work type.

|  | Front-line (%) | Backoffice (%) | Management (%) | Company (%) |
| --- | --- | --- | --- | --- |
| 18-49 | 80 | 50 | 20 | ~70 |
| 50-64 | 15 | 45 | 60 | ~25 |
| 65+ | 5 | 5 | 20 | ~5 |
| Overall Company | 70 | 25 | 5 |  |

**Table 2:** Total Savings in USD for Different Work Types and Vaccination Percentages.

| Work Type | Avg. Salary | 0% | 20% | 40% | 60% | 80% |
| --- | --- | --- | --- | --- | --- | --- |
| Front Line | \$43,421 | \$0 | \$258,352 | \$351,943 | \$461,277 | \$564,272 |
| Backoffice | \$43,421 | \$0 | \$85,494 | \$116,328 | \$151,114 | \$184,363 |
| Management | \$75,000 | \$0 | \$33,440 | \$46,783 | \$60,389 | \$76,405 |
| Company | \$45,000 | \$0 | \$377,286 | \$515,054 | \$672,780 | \$825,040 |

As illustrated, the total savings increase significantly with higher vaccination coverage. For front-line workers, total savings grow from $258,352 at 20% vaccination coverage to $564,272 at 80%. Back-office savings follow a similar pattern, rising from $85,494 to $184,363. Management sees more modest savings increases, from $33,440 to $76,405. Overall, the company’s total savings increase from $377,286 at 20% coverage to $825,040 at 80% coverage.

Additional to looking at the cost-benefit over the entire influenza season, the model forecasted timings of infections and peak infections and sickness (Figure 6.) At no and lower vaccine coverage, the peak in sickness across the company is significantly higher and occurs sooner, peaking in mid-January 2025, while at 80% vaccine coverage the peak in sickness is almost 3 times lower, with a delayed peak in mid-February 2025.

**Figure 6.**
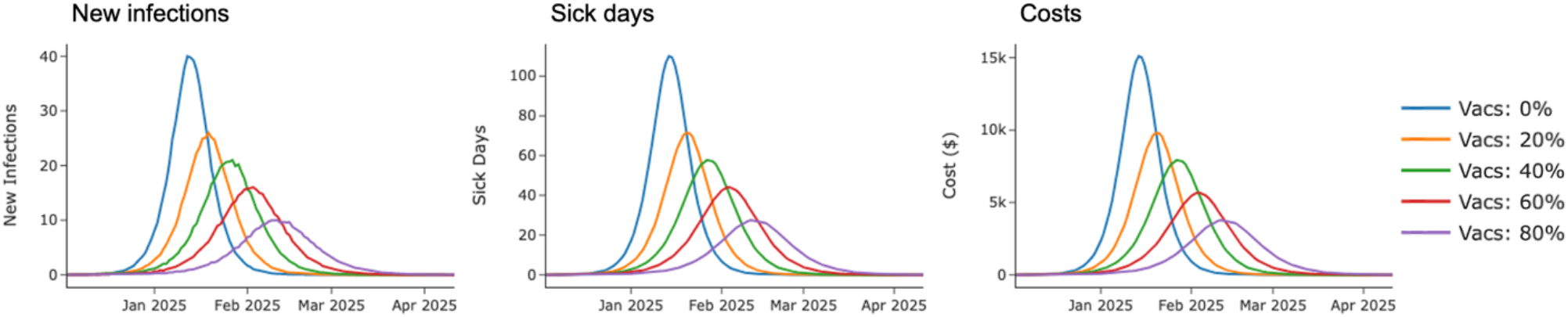
Plots showing forecasted scenario outcomes over time for different metrics across different vaccine coverage scenarios from 0% to 80% vaccine coverage.

**Figure 7.**
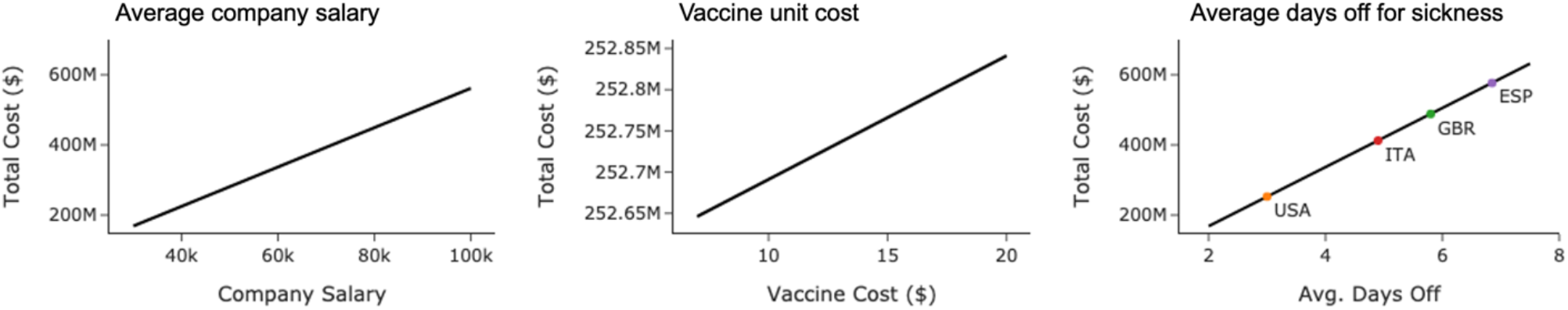
Plots showing the total cost for changes in the average company salary, the cost per vaccine and the average number of days off. The right-hand plot contains the costs associated with country specific average days off.

**Figure 8.**
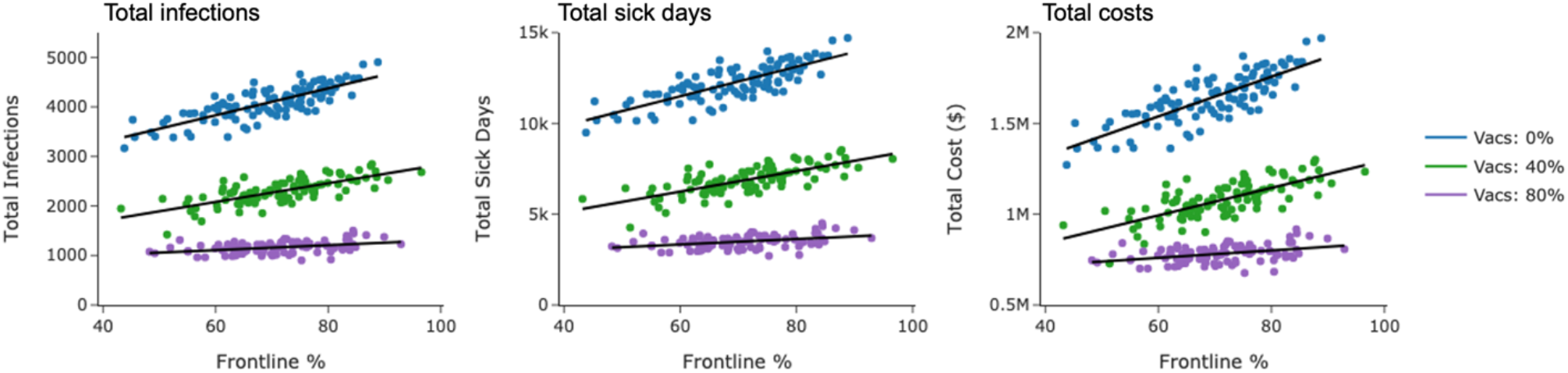
Plots showing the total infections, total sick days and the total cost for different proportions of the company being frontline workers, at three vaccination levels, (0%, 40% and 80%).

The results indicate that increasing vaccination coverage yields substantial financial benefits for the company. Total savings increase with higher vaccination rates, highlighting the cost-effectiveness of broader vaccination programs. Furthermore, the daily peak in sickness is significantly lower for higher vaccination coverage. The absolute financial impact is greater for frontline reflecting the increased number of workers in this group and the different infection dynamics across work types, while saving per capita are higher in the management group due to higher salaries.

### Sensitivity analysis

The sensitivity analysis shows that the total costs are significantly impacted by the average company salary and the average days off per infection, both of which can vary a lot depending on industry and the company’s country. With costs due to influenza potentially costing a company Spain over twice as much as the same company if it were based in the United States, due to employees in Europe tending to take more sick days than in the US [18].

The proportion of the company that are frontline workers also have significant impacts of the total sick days and the overall cost due to sickness, due to their increased contact with the general public. Increased vaccine coverage also reduces the total level of sickness more in companies with a higher proportion of frontline workers.

## Discussion

In this study we developed a novel machine learning-based SIR agent-based model to assess the cost-benefit of influenza vaccinations within workforces that employ significant proportions of frontline workers, such as public transport companies and hospitals. Our model provides an effective scenario analysis tool for calculating potential cost savings from workplace vaccination programs. This capability is essential for organisations aiming to evaluate the economic impact of different vaccination coverage levels. Previous studies have demonstrated the importance of modelling in assessing vaccination programs’ cost-effectiveness. For example, a study by Prosser et al. underscores the utility of detailed economic modelling in evaluating influenza vaccination strategies and their impact on public health [19]. Similarly, the work of Postma et al. highlights how dynamic models can aid in understanding the financial implications of vaccination interventions in different settings [20, 21] [21].

The machine learning agent-based SIR model used in this study offers several advantages over traditional vaccination models. Traditional deterministic models often use static parameters and simpler assumptions, which can limit their accuracy. For example, a study by Alexander et al. highlighted the limitations of static models in capturing the complexities of influenza transmission dynamics and the benefits of using more dynamic approaches [22]. Our model, by incorporating time-varying parameters and detailed sub-populations, provides a more nuanced view of influenza impacts, akin to the improvements noted in more sophisticated models such as those by Shaman et al., which integrated real-world data for better predictive accuracy [23].

The dynamic machine learning agent-based SIR model produced here was used to evaluate the financial benefits of influenza vaccination campaigns within frontline companies, revealing significant potential savings for organisations with higher vaccination coverage. The results showed that companies benefit most from vaccinations when they have higher average salaries and greater absenteeism rates. Moreover, organisations with a larger proportion of frontline workers experience more substantial benefits from vaccination programs. These findings are supported by research indicating that sectors with higher wages and absenteeism face greater economic impacts from influenza, as seen in our results with greater costs per person for the higher paid management group. For instance, the study by [7] demonstrated that workplace vaccination programs can be particularly cost-effective in high-wage sectors due to the higher costs associated with employee absenteeism [7]. Additionally, companies in regions with higher average sick leave, such as those in Europe, might see even greater cost savings. This observation is consistent with the study by [24] which reports that higher absenteeism rates in some countries lead to larger economic impacts from influenza [24]. An additional key finding from our model is that higher vaccination coverage results in a significantly lower peak in sickness levels. This has substantial implications for businesses, as it means there would not be a period of high sickness levels that would be difficult to manage and cover for. Reduced peak sickness levels can prevent operational disruptions and maintain productivity, which is crucial for business continuity. This benefit is supported by literature indicating that high vaccination coverage can flatten the epidemic curve, reducing the strain on healthcare systems and businesses alike [25, 5]. By avoiding peak periods of illness, businesses can manage absenteeism more effectively and reduce the need for temporary hires or overtime work, further contributing to cost savings and operational stability.

The strengths of our model include its detailed scenario analysis and use of real-world data, which better reflect real-world transmission dynamics and enable tailored vaccination strategies. The dynamic model allows it to be applied to more granular scenarios, and accounts for the dynamic nature of influenza. However, the linear relationship between vaccination rates and model parameters may oversimplify extreme scenarios. Future research could refine the model by incorporating non-linear effects and accounting for further external interactions to provide a more comprehensive analysis.

## Conclusion

The machine learning agent-based SIR model developed in this study offers a valuable tool for assessing the financial impact of influenza vaccination programs within the workplace. By providing detailed projections based on different vaccination scenarios, while accounting for differing work types and demographics, the model helps organisations make informed decisions about health interventions and budget allocations. The model’s ability to predict reduced peak sickness levels further underscores its utility in maintaining business continuity and operational efficiency.

## Data Availability

All data produced in the present study are available upon reasonable request to the authors

